# Quantitative assessment of the risk of airborne transmission of SARS-CoV-2 infection: prospective and retrospective applications

**DOI:** 10.1101/2020.06.01.20118984

**Authors:** G. Buonanno, L. Morawska, L. Stabile

**Author notes:** Corresponding author: Luca Stabile Department of Civil and Mechanical Engineering, University of Cassino and Southern Lazio, Cassino, FR, Italy Via G. Di Biasio 43, 03043 Cassino (FR), Italy.

## Abstract

Airborne transmission is a recognized pathway of contagion; however, it is rarely quantitatively evaluated. This study presents a novel approach for quantitative assessment of the individual infection risk of susceptible subjects exposed in indoor microenvironments in the presence of an asymptomatic infected SARS-CoV-2 subject. The approach allowed the maximum risk for an exposed healthy subject to be evaluated or, starting from an acceptable risk, the maximum exposure time. We applied the proposed approach to four distinct scenarios for a prospective assessment, highlighting that, in order to guarantee an acceptable individual risk of 10^−3^ for exposed subjects in naturally ventilated indoor environments, the exposure time should be shorter than 20 min. The proposed approach was used for retrospective assessment of documented outbreaks in a restaurant in Guangzhou (China) and at a choir rehearsal in Mount Vernon (USA), showing that, in both cases, the high attack rate values can be justified only assuming the airborne transmission as the main route of contagion. Moreover, we shown that such outbreaks are not caused by the rare presence of a superspreader, but can be likely explained by the co-existence of conditions, including emission and exposure parameters, leading to a highly probable event, which can be defined as a “superspreading event”.

## 1. Introduction

The airborne transmission of a virus and the consequent contagion risk assessment is a complex issue that requires multidisciplinary knowledge. It is necessary to understand the characteristics and mechanisms behind the generation of respiratory microdroplets^1,2^, the survival of viruses in microdroplets^3^, the transport of microdroplets and human exposure to them^4^, and the airflow patterns that carry microdroplets in buildings^5^. Expiratory human activities generate virus-carrying microdroplets that are small enough to remain aloft in air during exhalation, talking, and coughing^2,6,7^. Atomization occurs in the respiratory tract, and droplets are expelled at high speed during expiration^8,9^. Toques of liquid originating from different areas of the upper respiratory tract are drawn out from the surface and broken into droplets of different sizes^10^. The findings of early investigations^11–13^ served as a foundation for subsequent studies involving temporal and spatial visualization methods using high-speed cameras^14^, particle image velocimetry^8^ and, above all, increasingly accurate particle counters^6^, which have facilitated the detailed characterization and quantitation of droplets expelled during various forms of human respiratory exhalation flows. The issue of the viral load emitted, however, remained difficult to solve. In the past, backward calculation was used to estimate the emission of an infected subject based on retrospective assessments of infectious outbreaks only at the end of an epidemic^15–18^. This led to the definition of emission values for each virus regardless of the type of respiratory act and the metabolic activity of the infected subject. Recently, the authors presented an approach to evaluate the viral load emitted by infected individuals with a view to provide new predictive capacities, not currently available^19^. This approach, based on the oral viral load and the infectivity of the virus, takes into account the effect of other parameters such as inhalation rate, type of respiratory activity, and activity level, to estimate the quanta emission rate. This value provides key information for engineers and indoor air quality experts to simulate airborne dispersion of diseases in indoor environments. Indeed, the use of exposure risk models in closed environments^20,21^ makes it possible to estimate contagion starting from the emission values of a contagious subject.

The overall approach of emission and exposure modelling represents an essential tool to be applied in enclosed spaces, and can support air quality experts and epidemiologists in the management of indoor environments during an epidemic for both prospective and retrospective assessments.

In this paper we apply a novel approach that takes into account the characteristics of the emitting subject, the microenvironment, and the exposed subject to calculate the probability of infection and the individual risk, for both prospective and retrospective assessments of airborne infectious transmission of SARS-CoV-2. In the case of prospective assessment, various exposure scenarios in indoor environments were analyzed in order to assess the influence of risk mitigation parameters.

In the case of retrospective assessment, we estimated the probability of infection and the individual risk of two documented outbreaks.

## 2 Materials and methods

To evaluate both prospective and retrospective assessments of the airborne transmission of SARS-CoV-2, we used a four-step approach to quantify the probability of infection, i.e. the ratio between infected cases and the exposed population due to exposure in a microenvironment where a SARS-CoV-2 infected subject is present. The four steps of the proposed approach are: i) evaluation of the quanta emission rate; ii) evaluation of the exposure to quanta concentration in the microenvironment; iii) evaluation of the dose of quanta received by an exposed susceptible subject; and iv) estimation of the probability of infection on the basis of a dose-response model. The simulations of the probability of airborne transmission of SARS-CoV-2 were performed applying a Monte Carlo method^22^. Further they adopted the infection risk assessment typically implemented to evaluate the transmission dynamics of infectious diseases and to predict the risk of these diseases to the public^17,20,21^.

Once the probability of infection was obtained, an approach to evaluate the individual infection risk, i.e. a parameter that also takes into account how likely the probability of infection can occur, was also implemented. Individual risk can be easily compared to an acceptable risk, i.e. a target reference risk that could be suggested by agencies and regulatory authorities to control the pandemic. In the following sections, the methodologies adopted to evaluate the probability of infection based on the four step approach (section 2.1) and the individual infection risk (section 2.2) are described. The application of the proposed approach for prospective and retrospective assessments is described in sections 2.3 and 2.4.

### 2.1 Estimation of the probability of infection

#### 2.1.1 Evaluation of the quanta emission rate: the forward emission approach

Recently, Buonanno et al.^19^ proposed a forward emission approach to estimate the quanta emission rate of an infectious subject on the basis of the viral load in the sputum and the concentration of droplets expired during different activities. A quantum is defined as the dose of airborne droplet nuclei required to infect a susceptible person. The quanta emission rate (ER_q_, quanta h^−1^) was evaluated as:

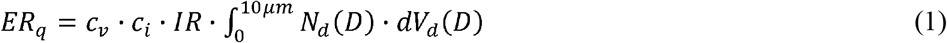

where *c*_v_ is the viral load in the sputum (RNA copies mL^-1^), *c*_i_ is a conversion factor defined as the ratio between one infectious quantum and the infectious dose expressed in viral RNA copies, *IR* is the inhalation rate (m^3^ h^-1^), *N*_d_ is the droplet number concentration (part. cm^−3^), and *V*_d_(*D*) is the volume of a single droplet (mL) as a function of the droplet diameter (*D*). The number and volume of the droplets (*V*_d_) is determined on the basis of data obtained experimentally by Morawska et al. (2009)^6^.

With reference to the SARS-CoV-2 viral load in the mouth, researchers have recently found c_v_, values of up to 10^11^ copies mL^−1^, which is also variable in the same patient during the course of the disease^23^–^26^. In particular, Rothe et al.^24^ reported a case of SARS-CoV-2 infection in which transmission appears to have occurred during the incubation period in the index patient. A high viral load of 10^8^ copies mL^−1^ was found, confirming that asymptomatic persons are potential sources of SARS-CoV-2 infection. Furthermore, Pan et al.^23^, in a study on 82 SARS-CoV-2 infected patients, found c_v_ values in the range of 10^8^−10^9^ RNA copies mL^−1^, also in the previous days and in the first days of onset of the disease. Consequently, the concentrations of the viral load in the mouth can reach values of 10 RNA copies mL^−1^ and occasionally up to 10 RNA copies mL^−1^. during the course of the disease.

The conversion factor, c_i_, i.e. the ratio between one infectious quantum and the infectious dose expressed in viral RNA copies, barely represents the probability of a pathogen surviving inside the host to initiate the infection; thus c_i_=1 implicitly assumes that infection will occur for each pathogen (RNA copy in the case of SARS-CoV-2) received by the exposed people. There are currently no values available in the scientific literature for c_i_ for SARS-CoV-2. Watanabe et al.^27^ estimated the infectious doses of several coronaviruses on the basis of data sets challenging humans with virus HCoV-229E (known as an agent of human common cold) and animals with other viruses (e.g. mice with MHV-1, considered as a surrogate of SARS-CoV-1). On the basis of the orders of magnitude of the infectivity conversion factors for the overall data sets, we assumed a c_i_ range between 0.01 and 0.1.

The quanta emission rate calculation was performed for four different emission profiles (which are adopted in the risk evaluations described later) evaluated as a combination of expiratory activities and activity levels: (i) oral breathing during resting; (ii) oral breathing during heavy activity; (ii) speaking during light activity; and (iv) singing (or loudly speaking) during light activity.

Quanta emission rates were calculated using eq. (1) and applying a Monte Carlo method^22^ in order to take into account for the possible variation of the input data. To this end, probability density functions characteristics of each parameter were considered. In particular, we considered normal distributions for: (i) the log-transformed c_v_ data (average and standard deviation of log_10_(c_v_) equal to 8 and 0.7 log_10_ (RNA copies mL^−1^), respectively); and (ii) the infectious dose c_i_ (average and standard deviation equal to 0.025 and 0.125, respectively). A distribution of quanta emission rates (ER_q_), was obtained as a result of application of the Monte Carlo method (Figure 1), i.e. the probability density function of ER_q_ (pdf_q_).

#### 2.1.2 Evaluation of the exposure to quanta concentration

The second step in evaluating the probability of infection is evaluation of the quanta concentration to which a susceptible subject is exposed. The quanta concentration at time *t, n*(*t*), in an indoor environment is based on the quanta mass balance proposed by Gammaitoni and Nucci^20^, and can be evaluated as:

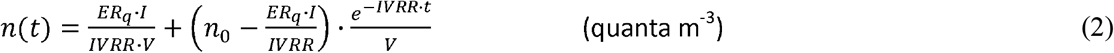

where *IVRR* (h^-1^) represents the infectious virus removal rate in the space investigated, *n_0_* represents the initial number of quanta in the space, *I* is the number of infectious subjects, *V* is the volume of the indoor environment considered, and ER_q_ is the quanta emission rate (quanta h^−1^) for the specific disease/virus under investigation. The quanta concentration calculation adopted here is based on the following hypotheses: the quanta emission rate is considered to be constant, the latent period of the disease is longer than the time scale of the model, and the droplets are instantaneously and evenly distributed in the room^20^. The infectious virus removal rate is the sum of three contributions^28^: the air exchange rate (AER) via ventilation, the particle deposition on surfaces (*k*, e.g. via gravitational settling), and the viral inactivation (*λ*). The deposition rate was evaluated as the ratio between the settling velocity of super-micrometric particles [roughly 1.0 × 10^−4^ m s^−1^ as measured by Chatoutsidou and Lazaridis^29^] and the height of the emission source (1.5 m); thus, *k* was 0.24 h^−1^. The viral inactivation was evaluated on the basis of the SARS-CoV-2 half-life (1.1 h) detected by van Doremalen et al.^3^, thus λ was 0.63 h^−1^.

In the exposure scenarios tested with the prospective and retrospective approaches, to take the variability of the input parameters into account, the indoor quanta concentration *n*(t) was determined through eq. (2), applying a Monte Carlo method that adopted the probability density functions (pdf_q_) characteristic of quanta emission rates (ER_q_). Since the probability density functions of the log-transformed log_10_(ER_q_) for the different expiratory activities resulted in a normal distribution (Shapiro-Wilk test, *p* < 0.01), the quanta concentration *n*(*t*) was evaluated by providing a Gaussian distribution of log_10_(ER_q_) (average and standard deviation values are is summarized in the results section; see Table 2) and then applying a back-transformation from log_10_(ER_q_) to ER_q_. The relative frequency at which a certain quanta concentration occurred for each time step of simulation, i.e. the probability density function of the quanta concentration (pdf_n_), was also obtained as result of the Monte Carlo simulations.

#### 2.1.3 Evaluation of the dose of quanta received by an exposed susceptible subject

The dose of quanta received by a susceptible subject exposed to a certain quanta concentration, *n*(*t*), for a certain time interval, *T*, can be evaluated by integrating the quanta concentration over time as:

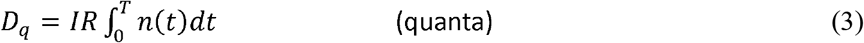

It can be concluded from Eq. (3) that the dose of quanta received by a susceptible subject is affected by the inhalation rate (*IR*) and subsequently by their activity level. As an example, for the same exposure scenario [i.e. identical *n*(*t*) and *T*], the dose of quanta received by subjects performing at a light activity level (IR = 1.38 m^3^ h^−1^; e.g. slowly walking) is more than double that received by people just sitting or standing (IR = 0.54 m^3^ h^−1^). For the dose, in the exposure scenarios described in this paper, the Monte Carlo method was applied to eq. (3) considering the probability density function of the quanta concentration (pdf_n_), whereas the IR was considered as a constant value; thus, the probability density function of the dose (pdf_D_) was obtained for each time step of the simulation.

#### 2.1.4 Evaluation of the probability of infection through a dose-response model

The fourth and final step in evaluating the probability of infection is the adoption of a dose-response model. Several dose-response models are available in the scientific literature for assessing the probability of infection of airborne-transmissible pathogens^16,17^, including deterministic and stochastic models, and threshold and non-threshold models.

The best-suited dose-response models for airborne transmission of pathogens are the stochastic models. In particular, exponential models have been mostly adopted in previous studies because of their suitability and simplicity^27^. Such models consider the pathogens as discrete bundles (i.e. quanta) distributed in a medium (e.g. saliva/sputum) in a random manner described by the Poisson probability distribution. When the medium is aerosolized, the pathogen distribution in the aerosols, and hence their distribution in the air, also follows the Poisson probability distribution. The complex Poisson summation equations can be simplified in an exponential equation^17,27,30^, i.e. the exponential dose-response model, which evaluates the probability of infection, P_I_ (%), of susceptible people as:

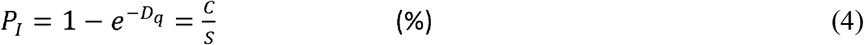

For a unit dose of quanta (*D_q_ =* 1), the probability of infection P_I_ is equal to 63%, from which derives the definition of “quantum” as the “amount of infectious material to infect 1-e^1^ (i.e. 63%) of the people in an enclosed space”^13,20^.

In the exponential dose-response model, the variation of host sensitivity to the pathogen is not considered. More complex models, such as the Beta-Poisson probability distribution, could take this factor into account^17,27,30^; nonetheless, in the present paper the differences in the exposed population in terms of susceptibility to the virus will not be considered.

The probability of infection P_I_ evaluated in the following exposure scenarios was determined through eq. (4), also applying a Monte Carlo method. To this end, the probability density functions of the dose of quanta (pdf_D_) obtained as a result of the Monte Carlo simulation on *D_q_* were considered; thus, a probability density function of P_I_ was also obtained (pdf_P_).

The probability of infection represents the ratio between the number of infection cases (C) and the number of exposed susceptibles (S). In retrospective analyses of documented outbreaks, the known C/S ratio is typically defined as the “attack rate”.

### 2.2 The individual infection risk and the basic reproduction number

As stated above, the probability of infection (P_I_) is the expected number of infection cases in relation to the number of exposed susceptibles (C/S ratio). However, based on eqs. (2–4), such probability is strongly influenced by the probability density function of the dose (pdf_D_), which is influenced in turn by the probability density function of the quanta concentration (pdf_n_) and by the probability density function of the quanta emission rate (pdf_q_). In other words, for a given exposure scenario (microenvironment, ventilation, inhalation rate of the exposed subject, etc.) the probability of infection (*P_I_*) can assume different values on the basis of the rate of quanta emitted by the infected subject: the lower the quanta emission rates, the lower the probability of infection (since all the other parameters affecting the exposure were considered to be constant values). Thus, when evaluating the individual risk (R) of an exposed person, we should know both the probability of infection (P_I_) and the probability of occurrence of such a P_I_ value (P_P_). The latter is defined by the probability density function pdf_P_. Since the probability of infection (P_I_) and the probability of occurrence P_P_ are independent events, the individual infection risk, R, can be evaluated as the product of the two terms:

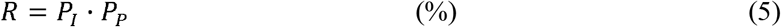

The probability density function of the individual risk, pdf_R_, can be obtained by multiplying all the possible P_I_ values obtained from the application of the Monte Carlo method to the four-step approach by the corresponding probability of occurrence. The maximum value of R in eq. (5), i.e. the mode of the pdf_R_, is of particular interest because it represents the most probable individual risk for a healthy subject or, in other words, the highest probability of being infected. In a conservative application of the proposed approach to estimate and reduce the risk of individuals being together with an infected individual in an indoor environment, the maximum individual infection risk must be less than an acceptable risk.

The US Environmental Protection Agency (EPA) typically uses a target reference risk range of 10^−4^to 10^−6^ for carcinogens in drinking water^31^, which is in line with World Health Organization (WHO) guidelines for drinking water quality, which base guideline values for genotoxic carcinogens on the upper bound estimate of an excess lifetime cancer risk of 10^−5^^32^. If the estimated lifetime cancer risk is lower than 10^−6^, the risk is considered acceptable, while risks above 10^−4^ are considered unacceptable^33^.

The choice of an acceptable contagion risk for SARS-CoV-2 is difficult and certainly questionable. However, considering the mortality rate of SARS-CoV-2, this turns out to be an order of magnitude lower than the corresponding value associated with carcinogenic diseases. For this reason, only for discussion purposes, the value of 10^−3^ is taken as an acceptable risk reference for SARS-CoV-2.

For the purpose of managing an epidemic and keeping the infection under control, it is also important to estimate the basic reproduction number of the infection, R_0_, which is calculated as the ratio between the number of susceptible people infected (C) and the infected subject (*I*). Thus, R_0_ can be easily evaluated by multiplying the infection probability, *P_I_*, by the number of exposed susceptible individuals (*S*). To control an epidemic, the R_0_ value must be less than **1**. Therefore, in addition to estimating an acceptable individual infection risk, it is necessary to specifically verify that, with the crowding expected within the environment, the corresponding value of R_0_ is less than 1.

### 2.3 Scenarios in the prospective assessment

The proposed four-step approach was applied to different indoor microenvironments by varying the main parameters in order to evaluate the effect of the influencing parameters. In particular, four emission profiles of the infected subject^6^ and corresponding profiles of the healthy subjects exposed were chosen. For the sake of simplicity, the simulations were run assuming that the susceptible subjects remained in the microenvironment for the same length of time as the infected subject (i.e. the two subjects enter and leave the environment under test together). Each indoor environment under investigation was tested for three different values of air exchange rate (AER). Table 1 presents a detailed summary of the four different indoor exposure scenarios considered to evaluate the risk of airborne transmission of SARS-CoV-2. Scenario A consists of a hospital room of 100 m^3^ where a resting infected patient emits quanta in the room through oral breathing, whereas the exposed susceptible subjects consist of a member of the medical staff in a light exercise activity (scenario A-1) and another patient at rest (scenario A-2). In scenario B, the infection affects two subjects, both oral breathing during a sports activity in a 300 m^3^ gym. Scenario C concerns two subjects (infected and healthy) in light activity while speaking in a generic 300 m^3^ office (bank, post office, supermarket, shop, etc.). Finally, scenario D represents an infected subject singing or speaking loudly in an 800 m^3^ room with healthy subjects listening at a sedentary activity level.

**Table 1.** Description of the exposure scenarios tested in the prospective assessment.

| Scenario A | Scenario B | Scenario C | Scenario D |
| --- | --- | --- | --- |

| Type of indoor environment | Hospital room | Gym | Public indoor environments<br>(e.g. restaurant, bank) | Conference room or auditorium |
| --- | --- | --- | --- | --- |
| Emitting subject | Patient<br>(Resting, oral breathing) | Exercising person<br>(heavy exercise, oral breathing) | Speaking person<br>(light exercise, voiced counting) | Singer or conference<br>loud speaker<br>(light exercise, unmodulated vocalization) |
| Exposed subject | A-1. Medical staff<br>(light exercise)<br>A-2. Patient (resting) | Exercising person<br>(heavy exercise) | Speaking person<br>(light exercise) | Spectator<br>(sedentary activity) |
| Volume ($\text{m}^3$) | 100 | 300 | 300 | 800 |
| Ventilation, AER ($\text{h}^{-1}$) | Natural ventilation $0.5 \text{ h}^{-1}$,<br>Mechanical ventilation $3 \text{ h}^{-1}$,<br>Mechanical ventilation $10 \text{ h}^{-1}$ | | | |
| Deposition rate, $k$ ($\text{h}^{-1}$) | 0.24 | | | |
| Inactivation rate, $\lambda$ ($\text{h}^{-1}$) | 0.63 | | | |

### 2.4 Retrospective assessments: outbreaks in a restaurant in Guangzhou, China, and at choir rehearsal in Skagit Valley (USA)

#### 2.4.1 The outbreak in a restaurant in Guangzhou, China

A possible case of airborne transmission was recently documented by Lu et al.^34^. Here, an index case patient traveled from the Chinese epidemic epicenter, Wuhan, on 23 January 2020 and ate lunch in a restaurant in Guangzhou, China, with his family on 24 January 2020 (family A, 10 people sitting at the same table). Later that day, the index patient experienced onset of fever and cough and SARS-CoV-2 infection was diagnosed. On the following days, nine other people were diagnosed with SARS-CoV-2 infection: four members from family A’s table and five other people at two different tables (families B and C). No other customers seated at other tables or waiters were infected.

The restaurant is a 5-floor building without windows; each floor has its own air ventilation system. The third floor dining area, at which the index patient ate lunch, has a floor area of 145 m^2^, with 15 tables arranged with a distance between each table of about 1 m. A total of 91 people (83 customers, 8 staff members) were in the room during that lunch. The exposure time was variable for the customers: those seated at tables close to the index patient had exposure times of 53 minutes (family B) and 73 minutes (family C). The ventilation and air conditioning situation is reported in Lu et al.^34^. Five fan coil air-conditioning units are installed in the room and there is no outdoor air supply; thus, the ventilation relies only upon infiltration and natural ventilation. The authors performed computational fluid dynamics analyses and tracer gas decay tests to obtain more information about the possible air-flow pathway in the room, and to determine the air exchange rate expected during that lunch. The analyses performed showed that, due to the particular installation and use of the fan coils, the room can be divided into different air-flow zones, with well-mixed conditions. The air-flow zone involving the table at which the index patient sat also included the two tables at which the other five infected people sat; and covered an area of roughly 45 m^3^. The tracer gas decay tests revealed a low air exchange rate (mostly due to the absence of an outdoor air supply) in the range of 0.56-0.77 h^−1^.

Therefore, on the basis of the available information, the retrospective assessment was applied to this outbreak case, through eqs. (2) and (3), using the following input data: i) room volume of 45 m^3^; ii) documented probability of infection, i.e. attack rate, of 45% (i.e. 5 out of 11 people of families B and C (family A members were excluded as they could easily have been infected through other infection routes); iii) average exposure time of 1 h; iv) speaking at a light activity level for all people (both emitting and exposed subjects), and v) average AER = 0.67 h^−1^.

#### 2.4.2 The outbreak at a choir rehearsal in Skagit (USA)

A further possible case of airborne transmission of SARS-CoV-2 was documented by the USA media (http://www.latimes.com/world-nation/story/2020-03-29/coronavirus-choir-outbreak). This case was recorded on 10 March, in Mount Vernon (Skagit County, Washington State, USA). In a 810 m^3^ hall, 61 choir members (out of a total of 121 regular members) gathered to rehearse, aware of the practices for the containment of contagion (frequent hand washing and social distancing). None of the members that attended had evident symptoms of SARS-CoV-2 infection. There was hand sanitizer at the front door and members refrained from the usual hugs and handshakes; each person brought their own sheet music. The event lasted from 6:30 pm to 9:00 pm (about 2.5 hours). Within few days, 33 of the 61 participants (53%) were diagnosed with SARS-CoV-2 infection, at least three were hospitalized, and two died^35^.

As pointed out by Hamner et al.^35^, the 2.5-hour singing practice could have provided several opportunities for droplet and fomite transmission (e.g. members sitting close to one another, sharing snacks, and stacking chairs at the end of the practice). Nonetheless, the abovementioned voluntary measures put in place would not support the documented spread of the contagion. On the contrary, the act of singing, itself, might have contributed to transmission through emission of aerosols, which is affected by loudness of vocalization^19^. This is even more relevant considering that attack rate of 53.3% (based on 33 confirmed cases) could represent a conservative estimate, since other 20 probable cases were mentioned by Hamner et al.^35^.

As regard the heating and ventilating system, limited information is available: the Fellowship Hall is heated by a relatively new commercial forced-air furnace with supply and return air grills situated high on a single wall. The furnace is installed to have both make-up and combustion air, but it is not known how much fresh air was provided on that evening. During the entire rehearsal no exterior doors were open. We applied a retrospective assessment to the case of the Skagit Valley choir through eqs. (2) and (3), using the following input data: i) room volume of 810 m^3^; ii) documented probability of infection, i.e. attack rate, equal to 53%; iii) exposure time of 2.5 h; iv) singing at a light activity level for all people; and v) natural ventilation with an AER = 0.5 h^−1^.

## 3 Results and Discussions

### 3.1 Statistics of quanta emission rates

Figure 1 and Table 2 show the statistics relating to the quanta emission rates for the four emission profiles considered in section 2.1. As shown in Buonanno et al.^19^, there are large differences between the emission profiles. Obviously the lowest values are found under the oral breathing condition during resting (median value of 0.36 quanta h^−1^), followed by the oral breathing condition during heavy activity as the inhalation rate increases (2.4 quanta h^−1^), and reaching 4.9 quanta h^−1^ for the increase in aerosol emitted during vocalization^6^ and, finally, peaking during singing/speaking loudly (31 quanta h^−1^). Indeed, the rate of particle emission during normal human speech is positively correlated with the amplitude of vocalization^36^.

**Figure 1.**
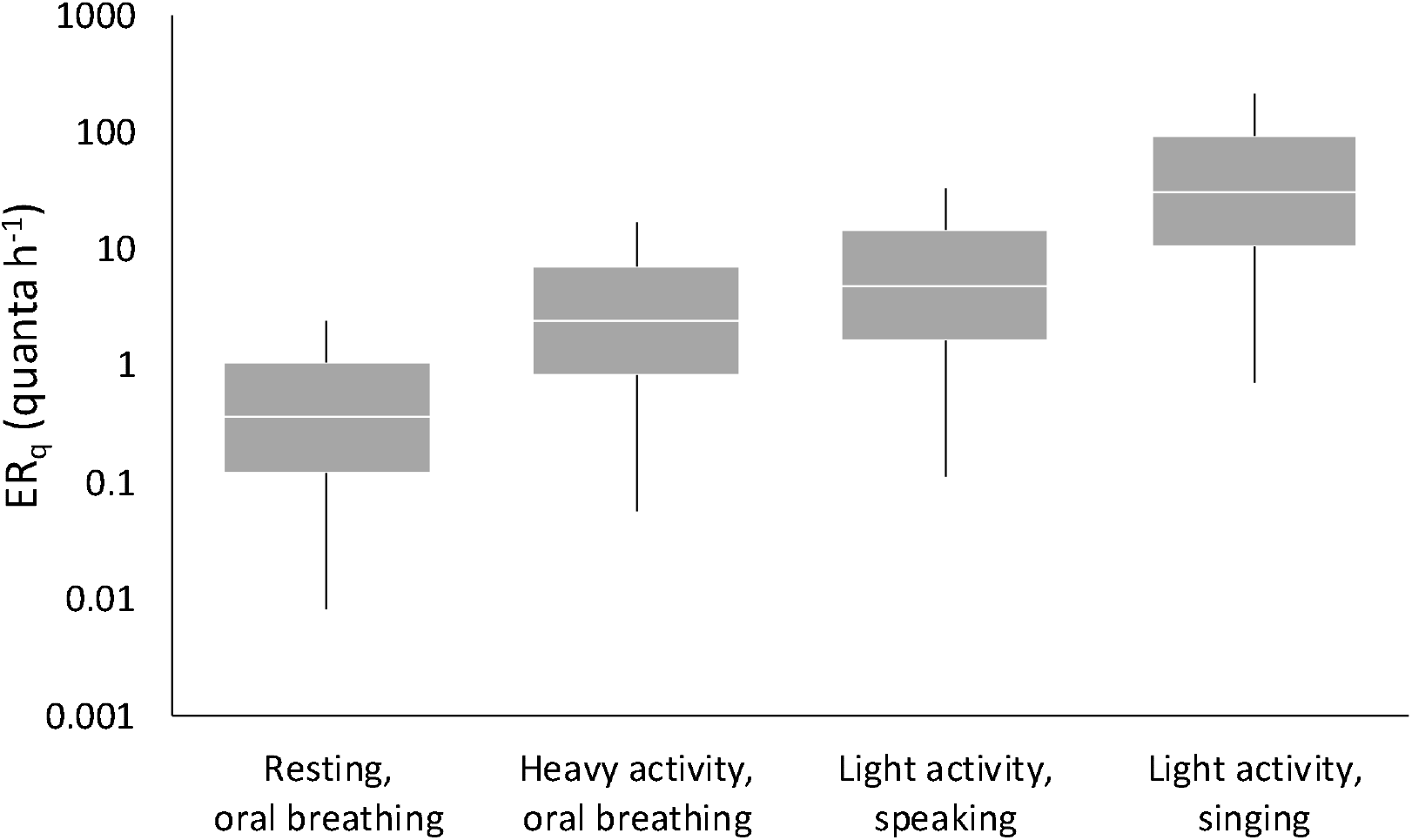
Statistics of quanta emission rates (ER_q_) for the four expiratory activities considered in the exposure scenarios. Data reported represent 1^st^, 25^th^, 50^th^, 75^th^, and 99^th^ percentiles.

The probability density functions of the quanta emission rates (P_q_) were also determined. In particular, the log-transformed ER_q_ values obtained from the Monte Carlo simulations resulted in a normal distribution (Shapiro-Wilk test, *p* < 0.01). Table 2 shows the average and standard deviation values of the log_10_(ER_q_).

**Table 2.** ER_q_ (quanta h^−1^) and log(ER_q_) statistics for SARS-CoV-2 as a function of the expiratory activity and activity level. The log-transformed ER_q_ values follow a log-normal distribution; thus, the average and standard deviation values of the log_10_(ER_q_) are provided.

|  |  | Resting,<br>oral breathing | Heavy activity,<br>oral breathing | Light activity,<br>speaking | Light activity,<br>singing (or speaking<br>loudly) |
| --- | --- | --- | --- | --- | --- |
| $ER_q$ | 5<sup>th</sup> percentile | $2.5 \times 10^{-2}$ | $1.7 \times 10^{-1}$ | $3.4 \times 10^{-1}$ | $2.1 \times 10^0$ |
| | 25<sup>th</sup> percentile | $1.2 \times 10^{-1}$ | $8.1 \times 10^{-1}$ | $1.6 \times 10^0$ | $1.0 \times 10^1$ |
| | 50<sup>th</sup> percentile | $3.6 \times 10^{-1}$ | $2.4 \times 10^0$ | $4.9 \times 10^0$ | $3.1 \times 10^1$ |
| | 75<sup>th</sup> percentile | $1.1 \times 10^0$ | $7.2 \times 10^0$ | $1.5 \times 10^1$ | $9.3 \times 10^1$ |
| | 95<sup>th</sup> percentile | $5.2 \times 10^0$ | $3.0 \times 10^1$ | $7.1 \times 10^1$ | $4.5 \times 10^2$ |
| | 99<sup>th</sup> percentile | $1.6 \times 10^1$ | $1.1 \times 10^2$ | $2.2 \times 10^2$ | $1.4 \times 10^3$ |
| $\log_{10}(ER_q)$ | Average | $-4.4 \times 10^{-1}$ | $3.9 \times 10^{-1}$ | $6.9 \times 10^{-1}$ | $1.5 \times 10^0$ |
| | Stand. dev | $7.1 \times 10^{-1}$ | $7.1 \times 10^{-1}$ | $7.1 \times 10^{-1}$ | $7.1 \times 10^{-1}$ |

We point out that the estimated values present two main uncertainty contributions clearly related to the limited data currently available for the SARS-CoV-2: i) a still low number of experimental data for the viral load in the mouth, c_v_, of SARS-CoV-2 infected subjects, ii) unavailable infectivity conversion factors, c_i_, for SARS-CoV-2; indeed, as mentioned in the methodology section, the c_i_ parameter was estimated on the basis of data available for other coronaviruses challenging humans (only in the case of HCoV-229E) and animals (for all other types of coronavirus).

### 3.2 Risk management in prospective assessment applications

#### 3.2.1 Illustrative example of probability of infection and individual risk evaluation

In Figure 2 an illustrative example of quanta concentration *n*(*t*), dose of quanta (D_q_), and probability of infection (P_<u>I</u>_) trends as a function of time (here shown for 2 h) resulting from the Monte Carlo simulation for exposure scenario D (singing exhibition, conference speaker) with an AER = 0.5 h^−1^ is shown. In particular, the trends of different percentiles are reported. The example shows that a person singing/speaking loudly in such a microenvironment can lead to a median *n*(*t*) value after 2 hours equal to 0.027 quanta h^−1^ (with a 5^th^-95^th^ percentile range of < 0.0020.38 quanta h^−1^). Such concentrations lead to a median dose of quanta received by the subject exposed for 2 h in a sedentary activity equal to 0.029 quanta (with a 5^th^-95^th^ percentile range of < 0.002-0.42 quanta), then resulting in a median probability of infection, P_I_, of 2.8% (with a 5^th^-95^th^ percentile range of 0.2%-33.0%). Thus, if higher quanta emission rates are considered, the indoor quanta concentrations and the consequent probability of infection can be more than 10 fold the median values.

**Figure 2.**
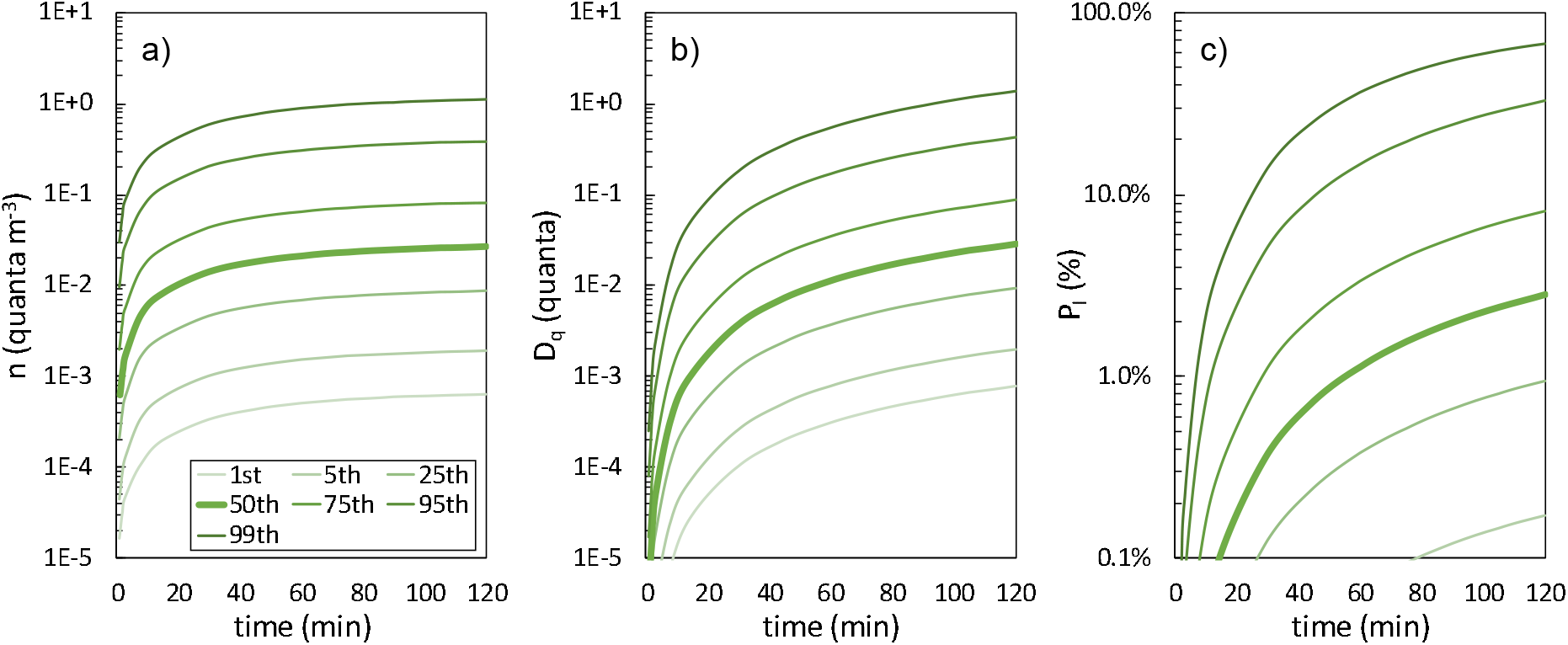
Trends of quanta concentration (a), dose of quanta (b), and probability of infection (c) as a function of time (here shown for 2 h of exposure) resulting from the Monte Carlo simulation for exposure scenario D with an AER = 0.5 h^−1^. Different percentiles are reported.

In view of the application of a conservative approach that could be essential to reduce the risk of contagion in indoor environments, using the highest quanta concentration and probability of infection values can be misguiding. Indeed, the probability of occurrence of such high values is extremely low. Thus, as described in section 2.2, a proper evaluation of the individual infection risk (R) can be obtained by applying eq. (4), i.e. multiplying the probability of infection (P_I_) by the corresponding probability of occurrence (P_P_). In Figure **3** the probability density functions of individual infection risk (pdf_R_), probability of infection (pdf_P_), quanta concentration (pdf_n_), and dose of quanta (pdf_D_) after 2 hours of exposure are reported (in terms of R, P_I_, n and D_q_ values for each percentile) for the illustrative example discussed above (i.e. scenario D, AER = 0.5 h^−1^). The individual infection risk (R) presents a maximum value (R_max_) at the 85^th^ percentile (2.2%) due to a probability of infection P_I_ = 14.5% and a probability of occurrence P_P_ = 15%. In other words, the R value at the 85^th^ percentile is the most probable individual infection risk for a healthy susceptible subject (i.e., the one with the highest chance of occurring). Due to the similarity of the probability density functions of the four expiration activities resulting from the calculation of the quanta emission rates (log_10_(ER_q_) reported in Table 2), the pdf_R_ for all the exposure scenarios tested here was similar to that of the exposure scenario shown in Figure 3 (i.e. the R_max_ value occurs in the narrow range of 84^th^-90^th^ percentile).

**Figure 3.**
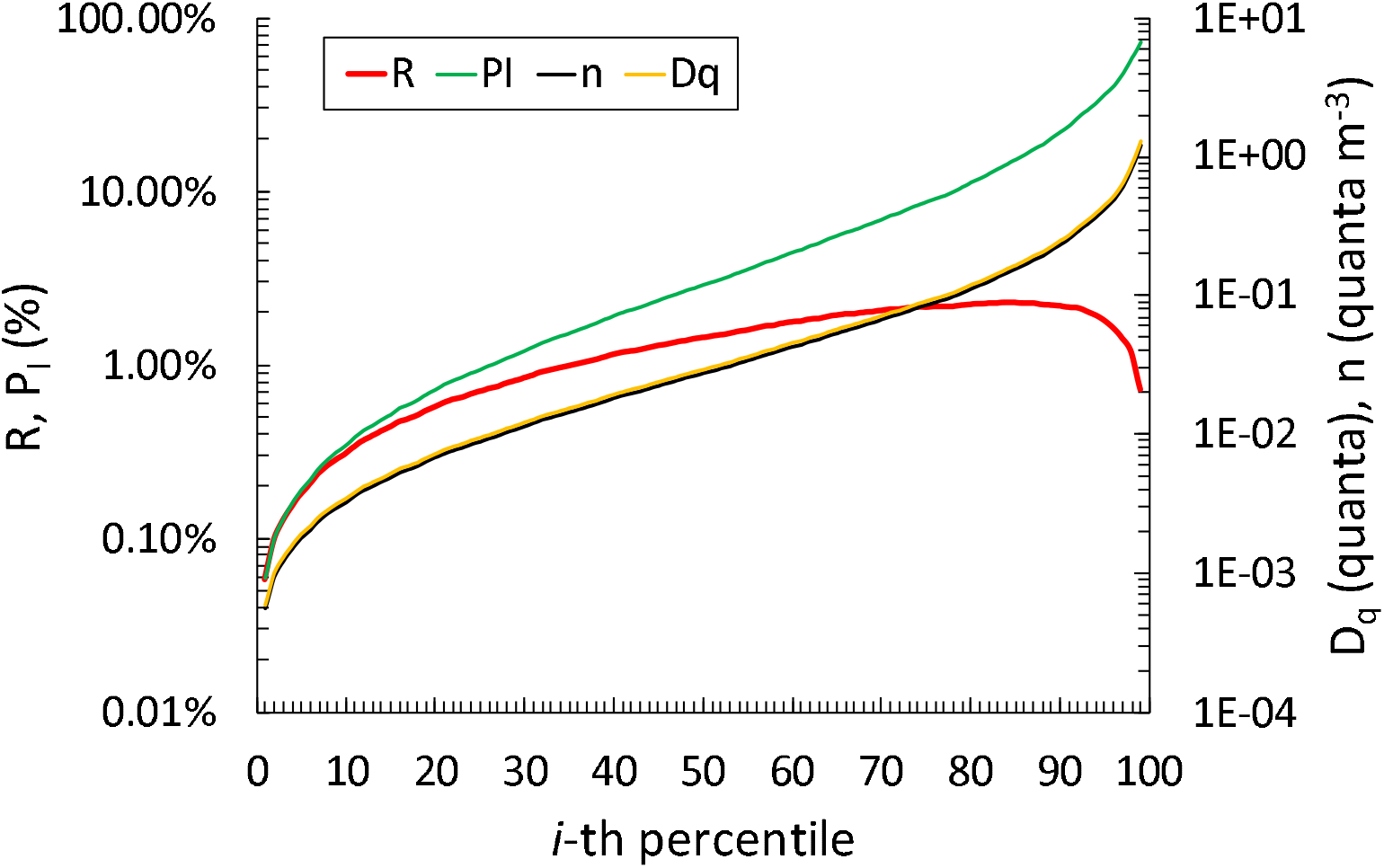
Probability density functions of individual infection risk, probability of infection, quanta concentration, and dose of quanta at t = 120 min for the illustrative example reported in Figure 2 (exposure scenario A with an air exchange rate of 0.5 h^−1^). The probability density functions are reported as quanta concentration (*n*), dose of quanta (D_q_), probability of infection (P_I_), and individual infection risk (R) for each percentile. The maximum individual infection risk (R_max_) is 1.9% and occurs at the 85^th^ percentile (P_I_ = 14.5%, P_q_ = 15%).

Furthermore, as discussed in section 2.2, the probability density function of the probability of infection (pdf_P_) is mostly influenced by the probability density function of the quanta emission rate (pdf_q_) when moving backwards in the four-step approach; indeed, once the exposure scenario is defined, all the parameters contributing to the calculation of P_I_ (ventilation, room volume, subject activity, etc.) can be considered as constant values. Thus, for a simplified estimate of R_max_, the simplest calculation can be applied (instead of the Monte Carlo method) by just adopting the 85^th^ percentile of the quanta emission rate in the four-step calculation using eqs. (2–4).

#### 3.2.2 Estimate of the maximum individual risk versus exposure time in indoor environments

Figure 4 and Table 3 show the results of the Monte Carlo simulations for the four exposure scenarios analyzed. The exposure time-risk relationships reported in Figure 4 are essential as they can be used by choosing either the exposure time or the maximum risk R_max_ as the independent variable. In the first case, knowing the exposure time of the healthy subject in the environment in question, the corresponding individual infection risk can be evaluated and then compared to an acceptable infection risk. In the second case, once an acceptable infection risk has been imposed, the corresponding maximum exposure time value can be easily assessed. The four scenarios are examined assuming an acceptable risk value of 10^−3^ as discussed in section 2.3. Since the maximum value of individual risk occurs roughly at the 90^th^ percentile, the corresponding probability of occurrence of the risk (P_P_) is 10%; thus, an acceptable individual infection risk of 10^−3^ will roughly correspond to a probability of infection of P_I_ = 1%. For indoor environments characterized by high crowding indexes a P_I_ < 1% is essential as it can assure a R_0_ < 1 when crowded with up to 100 people. Therefore, assuring an individual infection risk of 10^−3^ also guarantees the control of the epidemic with an R_0_ < 1 for a maximum number of exposed healthy people S < 100.

**Table 3.** Maximum exposure time (min) for the different exposure scenarios to reach an acceptable maximum individual infection risk (R_max_).

| Exposure scenarios | AER ($\text{h}^{-1}$) | Maximum individual infection risk ($R_{\max}$) | | | | |
| --- | --- | --- | --- | --- | --- | --- |
| | | $1 \times 10^{-1}$ | $1 \times 10^{-2}$ | $1 \times 10^{-3}$ | $1 \times 10^{-4}$ | $1 \times 10^{-5}$ |
| Scenario A-1 - Hospital room | 0.5 | 2797 | 212 | 36 | 10 | 3 |
| Emitting subject: patient | 3.0 | 7968 | 600 | 56 | 11 | 3 |
| Exposed subject: Medical staff | 10.0 | 22430 | 1727 | 157 | 17 | 3 |
| Scenario A-2 - Hospital room | 0.5 | 8039 | 597 | 72 | 17 | 5 |
| Emitting subject: patient | 3.0 | 21739 | 1678 | 159 | 21 | 5 |
| Exposed subject: patient | 10.0 | 64333 | 4671 | 455 | 46 | 6 |
| Scenario B – Gym | 0.5 | 519 | 55 | 14 | 4 | 1 |
| Emitting subject: Exercising person | 3.0 | 1500 | 110 | 16 | 4 | 1 |
| Exposed subject: Exercising person | 10.0 | 4119 | 314 | 29 | 5 | 1 |
| **Scenario C – Public indoors** | 0.5 | 627 | 60 | 15 | 5 | 1 |
| Emitting subject: Speaking person | 3.0 | 1812 | 137 | 19 | 5 | 1 |
| Exposed subject: Speaking person | 10.0 | 4807 | 372 | 36 | 6 | 1 |
| **Scenario D – Conference room** | 0.5 | 652 | 62 | 16 | 5 | 1 |
| Emitting subject: Singer | 3.0 | 1826 | 138 | 19 | 5 | 1 |
| Exposed subject: Spectator | 10.0 | 5187 | 392 | 38 | 6 | 2 |

**Figure 4.**
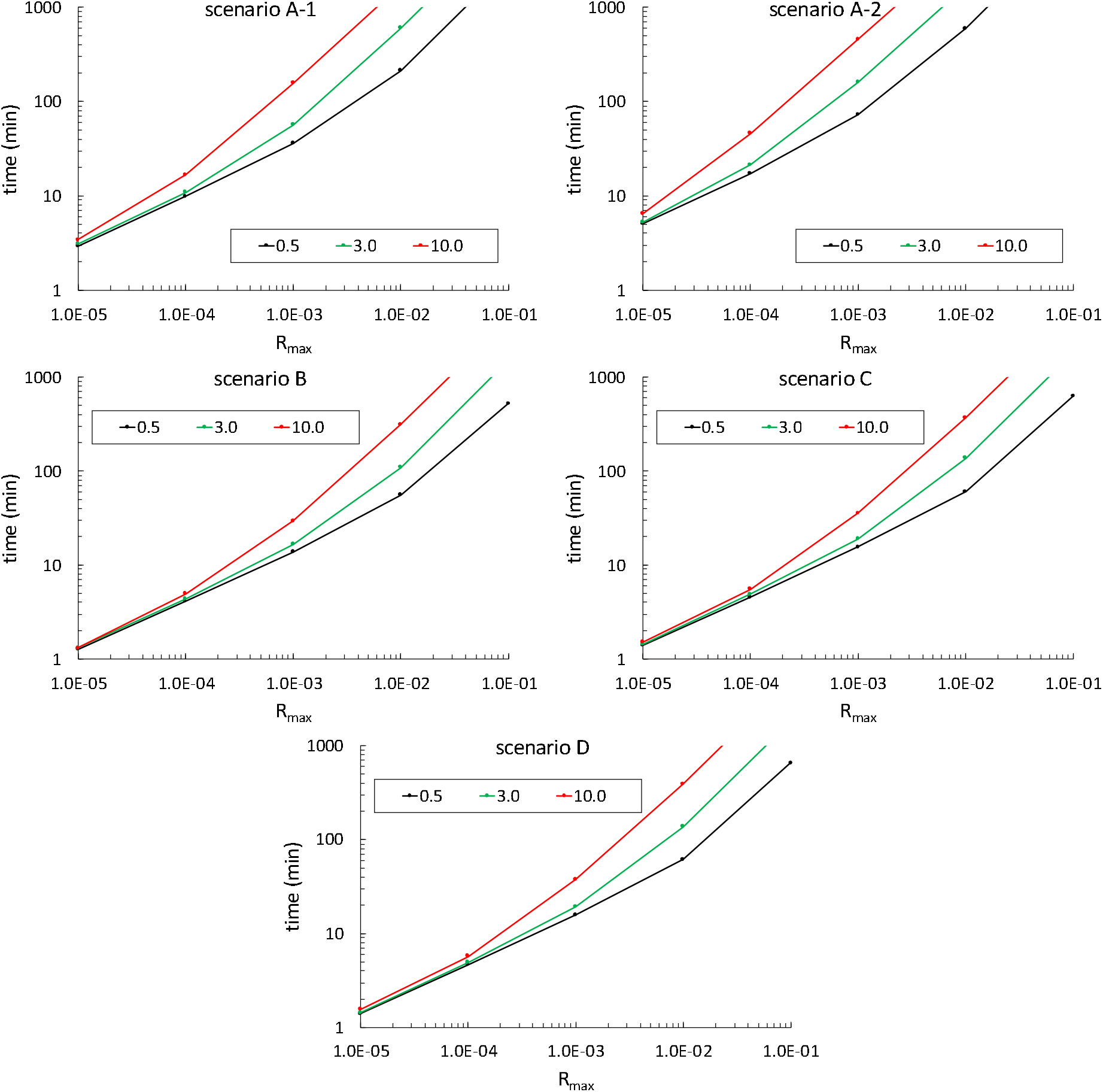
Relationship between time of exposure and individual risk (R) as a function of the air exchange rate (0.5 h’^1^, 3 h’^1^, and 10 h^−1^) for the exposure scenarios investigated in the prospective approach and summarized in Table 1.

For the exposure scenario discussed above (scenario D, AER = 0.5 h^−1^) the maximum exposure time to reach an accepted risk of R = 10^−3^ is very short (16 min); this is due to the high viral load emitted during singing or speaking loudly leading to high quanta concentrations despite the large volume available. Obviously, the exposure time can increase with higher ventilation rates, e.g. reaching 38 min in the case of mechanical ventilation at 10 h^−1^. The crowding index of such an indoor environment (800 m^3^) ranges from 0.75 m^2^ (auditorium) to 2 m^2^ (conference room) per person^37^; thus, for a room height of 4 m a corresponding floor area of 200 m^2^ will be available, then resulting in a total number of people simultaneously present in the room (S) ranging from 100 (conference room) to 267 (auditorium). Therefore, after 16 min of exposure in the case of natural ventilation (or 38 min in the case of mechanical ventilation with AER = 10 h^−1^), the value of R_0_ will be higher (auditorium) or equal (conference room) to 1. Thus, in the management of the epidemic, reducing the crowding index could be essential. Accepting higher R_max_ values would clearly increase the maximum exposure time; indeed, in the case of R_max_ = 10^−2^, the exposure time values would be 62 min and 392 min, for an AER equal to 0.5 h^−1^ and 10 h^−1^, respectively. However, in this case, the corresponding value of R_0_ would be lower than 1 only for a number of exposed subjects lower than 10.

In scenario C, the infected subject in light activity speaks in a 300 m^3^ environment, along with the healthy subject. The simultaneous reduction of both the quanta emission rate and the volume compared to scenario D makes the maximum exposure times for an acceptable infection risk of 10^−3^ comparable to the previous case (15 min and 36 min for ventilation of 0.5 h^−1^ and 10 h^−1^, respectively). Additionally, in this case, the estimated exposure times would guarantee an R_0_ < 1 with S < 100 subjects.

In scenario A (patient emitting at rest in oral breathing), the maximum exposure time in a hospital room of 100 m^3^ for both a medical staff member (scenario A-1) and a patient at rest without infection (scenario A-2) is evaluated. In both cases the exposure times increase significantly with the ventilation rate, reaching 36 min and 157 min (scenario A-1), and 72 min and 455 min (scenario A-2) with AER values of 0.5 h^−1^ and 10 h^−1^, respectively. However, despite the small size of the room, the ER_q_ was extremely small (Table 2); thus, unless a large number of infected subjects is simultaneously present in the room, the concentration of viral load in a hospital room can be considered low; nonetheless, the overall risk may become relevant due to the long exposure times (of 36 and 157 min). Finally, in exposure scenario B (the gym with infected and healthy subjects during heavy activity with oral breathing), although there is no vocalization in the subject’s activity, the high inhalation rate produces considerable ER_q_ values, then increasing the individual risk; thus, in order to guarantee an acceptable infection risk of 10^−3^ the maximum exposure times resulted quite short, i.e. 14 min and 29 min for 0.5 h^−1^ and 10 h^−1^, respectively.

Thus, for all the scenarios investigated, the ventilation conditions strongly influence the risk (or the exposure time) of the exposed subject: this difference increases as the accepted risk increases as shown in the trends presented in Figure 4. In contrast, if a lower risk was accepted (i.e. 10^−4^ or 10^−5^), increasing the air exchange rate is not leading to the significant reduction of the risk, and local exhaust ventilation would be more effective.

### 3.3 Retrospective assessment application: the outbreaks in a restaurant in Guangzhou and at a choir rehearsal in Skagit Valley

Figure 5 shows the trends of quanta concentration and probability of infection (P_I_) evaluated for the retrospective cases defined in section 2.4 (a restaurant in Guangzhou and the Skagit Valley choir). The retrospective analysis applied to the restaurant in Guangzhou (Figure 5a) revealed that, under the boundary conditions considered in the simulation (in terms of room volume, ventilation, number of exposed people; see section 2.4.1), a probability of infection (P_I_) after 1 hour of exposure equal to the attack rate (45%) can be reached for a quanta emission rate of ER_q_ = 61 quanta h^1^. This emission rate, for an emitting subject speaking during light exercise, occurs at the 93^rd^ percentile of the probability density function of ER_q_ (P_q_).

**Figure 5.**
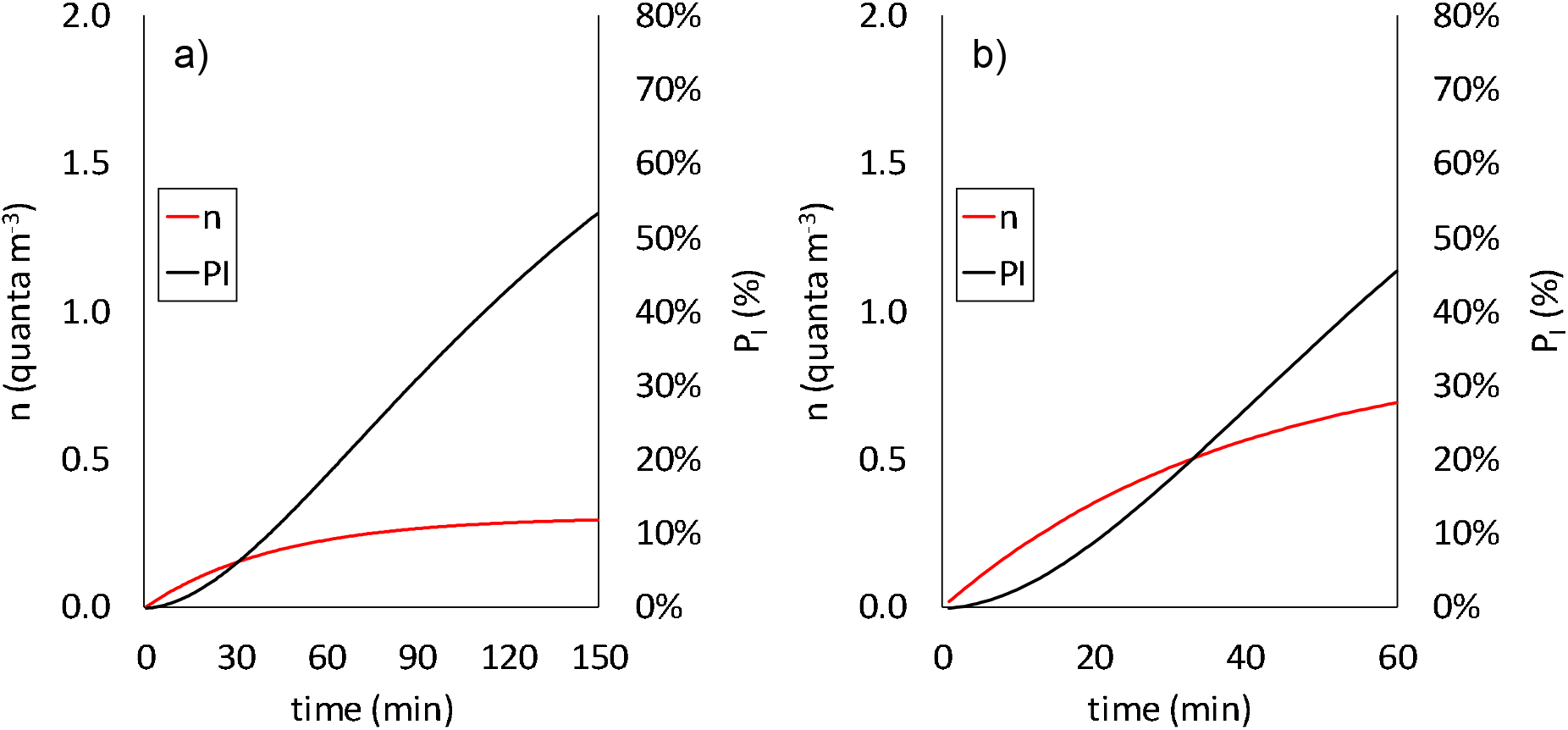
Quanta concentration (n) and probability of infection (P_I_) evaluated for the retrospective cases applied at the documented outbreaks at (a) the restaurant in Guangzhou and (b) the Skagit Valley choir.

Similarly, for the retrospective analysis applied to the Skagit Valley choir (Figure 5b), in order to reach an attack rate of 53% after 2.5 hours of exposure under the simulation boundary conditions reported in section 2.4.2, a quanta emission rate of 341 quanta h^−1^ is needed. Additionally, in this case, such an emission rate occurs at the 92^nd^ percentile of the probability density function (P_q_) of an infected subject while singing.

Therefore, for both the analyzed cases in the retrospective analyses, the required ERq values to obtain the documented R_e_ fall perfectly within the possible values of the emission profiles under consideration (i.e. speaking and singing/speaking loudly in light activity reported in Table 2). Moreover, such emission values incur high individual infection risks as they are around the 90^th^percentile, i.e. at the percentile maximizing the individual infection risk (R_max_). Indeed, the R values for the restaurant at Guangzhou and the Skagit Valley choir were 3.2% and 3.7%, respectively – more than one order of magnitude higher than the acceptable risk of 10^−3^. In these two cases, an individual risk of < 10^−3^ would have been obtained with a probability of infection P_I_ = 1.3-1.4%: such a P_I_ is not actually achievable by varying and optimizing the room ventilation (e.g. AER>100 h^−1^ would be required), and is achievable only by reducing the exposure time of the susceptible subjects and the quanta emission rates.

To summarize, the retrospective assessment of the two SARS-CoV-2 outbreaks investigated demonstrate that the documented number of infected people can be explained by means of the airborne transmission route; indeed, the most probable of the expected events occurred. The approach and consequent calculation reported here clearly highlights that the explanation of such a high number of infected people does not necessarily require the presence of a superspreader in the environment (i.e. an infected person with the highest viral load, C_v_, and infectious dose, c_i_), but rather a co-existence of conditions, including emission and exposure parameters, leading to a highly probable event, which can be defined as a “superspreading event”.

## Data Availability

We confirm that all the data are available

